# Evaluation of a Machine Learning Approach Utilizing Wearable Data for Prediction of SARS-CoV-2 Infection in Healthcare Workers

**DOI:** 10.1101/2021.11.04.21265931

**Authors:** Robert P. Hirten, Lewis Tomalin, Matteo Danieletto, Eddye Golden, Micol Zweig, Sparshdeep Kaur, Drew Helmus, Anthony Biello, Renata Pyzik, Erwin P Bottinger, Laurie Keefer, Dennis Charney, Girish N. Nadkarni, Mayte Suarez-Farinas, Zahi A. Fayad

**Author notes:** **Correspondence:** Robert P Hirten MD, 1468 Madison Avenue, Annenberg Building RM 5-12, New York, NY 10029. **Full Address of Authors:** Robert P Hirten MD, 1468 Madison Avenue, Annenberg Building RM 5-12, New York, NY 10029. Matteo Danielleto PhD, 770 Lexington Ave, New York, NY 10065 Lewis Tomalin PhD, 1425 Madison AveNew York, NY 10029, Icahn Building Floor 2nd Room L2-70C, Box 1077 Micol Zweig MPH, 770 Lexington Ave, New York, NY 10065 Eddye Golden MPH, 770 Lexington Ave, New York, NY 10065 Sparshdeep Kaur BBA, 770 Lexington Ave, New York, NY 10065 Drew Helmus MPH, 1468 Madison Avenue, Annenberg Building RM 5-12, New York, NY 10029. Anthony Biello BA, 1468 Madison Avenue, Annenberg Building RM 5-12, New York, NY 10029. Renata Pyrik MS, Leon and Norma Hess Center for Science and Medicine. 1470 Madison Avenue, 1st Floor, New York, NY 10029. Dennis Charney MD, 1468 Madison Ave, Annenberg Building Floor 21, New York, NY 10029 Erwin P Bottinger MD, 770 Lexington Ave, New York, NY 10065 Laurie Keefer PhD, 17 East 102nd Street, Box 1134, New York, NY 10029 Mayte Suarez-Farinas PhD, 1425 Madison AveNew York, NY 10029, Icahn Building Floor 2nd Room L2-70C, Box 1077 Girish N Nadkami MD, 770 Lexington Ave, New York, NY 10065 Zahi A Fayad PhD, Leon and Norma Hess Center for Science and Medicine. 1470 Madison Avenue, 1st Floor, New York, NY 10029. **Authors Contributions:** Author Contributions: RPH, GN, MD, ZAF developed the study concept. RPH and LT assisted with the drafting of the manuscript. RPH, LT, MD, EG, MZ, SK, DH, AB, RP, DC, EPP, LK, GNN, MSF, ZAF critically revised the manuscript for important intellectual content. RPH, LT, MD, EG, MZ, SK, DH, AB, RP, DC, EPP, LK, GNN, MSF, ZAF provided final approval of the version of the manuscript to be published and agree to be accountable for all aspects of the work. All authors approve the authorship list. All authors had full access to all the data in the manuscript and had final responsibility for the decision to submit for publication. RPH, ZAF, MSF, MD, and LT have verified the underlying data.

## Abstract

**Importance:** Passive and non-invasive identification of SARS-CoV-2 infection remains a challenge. Widespread use of wearable devices represents an opportunity to leverage physiological metrics and fill this knowledge gap.

**Objective:** To determine whether a machine learning model can detect SARS-CoV-2 infection from physiological metrics collected from wearable devices.

**Design:** A multicenter observational study enrolling health care workers with remote follow-up.

**Setting:** Seven hospitals from the Mount Sinai Health System in New York City

**Participants:** Eligibility criteria included health care workers who were ≥18 years, employees of one of the participating hospitals, with at least an iPhone series 6, and willing to wear an Apple Watch Series 4 or higher. We excluded participants with underlying autoimmune/inflammatory diseases, and medications known to interfere with autonomic function. We enrolled participants between April 29^th^, 2020, and March 2^nd^, 2021, and followed them for a median of 73 days (range, 3-253 days). Participants provided patient-reported outcome measures through a custom smartphone application and wore an Apple Watch, collecting heart rate variability and heart rate data, throughout the follow-up period.

**Exposure:** Participants were exposed to SARS-CoV-2 infection over time due to ongoing community spread.

**Main Outcome and Measure:** The primary outcome was SARS-CoV-2 infection, defined as ±7 days from a self-reported positive SARS-CoV-2 nasal PCR test.

**Results:** We enrolled 407 participants with 49 (12%) having a positive SARS-CoV-2 test during follow-up. We examined five machine-learning approaches and found that gradient-boosting machines (GBM) had the most favorable 10-CV performance. Across all testing sets, our GBM model predicted SARS-CoV-2 infection with an average area under the receiver operating characteristic (auROC)=85% (Confidence Interval 83-88%). The model was calibrated to improve sensitivity over specificity, achieving an average sensitivity of 76% (CI ±∼4%) and specificity of 84% (CI ±∼0.4%). The most important predictors included parameters describing the circadian HRV mean (MESOR) and peak-timing (acrophase), and age.

**Conclusions and Relevance:** We show that a tree-based ML algorithm applied to physiological metrics passively collected from a wearable device can identify and predict SARS-CoV2 infection. Utilizing physiological metrics from wearable devices may improve screening methods and infection tracking.

## Introduction

Infection prediction traditionally relies on the development of characteristic symptomatology, prompting confirmatory diagnostic testing. However, the SARS-CoV-2 infection poses a challenge to this traditional paradigm given its variable symptomatology, prolonged incubation period, high rate of asymptomatic infection, and variable access to testing.^1,2^ Ongoing case surges throughout the world, prompted by the delta variant, are characterized by greater infectivity and raise the possibility that SARS-CoV-2 may become endemic. While highly effective vaccines against SARS-CoV-2 have been developed, limited vaccine supplies, low vaccination rates in some communities and the evolution of variants, have prompted ongoing infectious spread.^3^ Novel means to identify and predict SARS-CoV-2 infection are needed.

Wearable devices are commonly used and can measure multi-modal continuous data throughout daily life.^4^ Increasingly, they have been applied to applications in health and disease.^5^ Researchers have previously demonstrated that the addition of wearable sensor data to symptom tracking apps can increase the ability to identify Corona Virus Disease-2019 (COVID-19) patients.^6^ Additionally, the combination of heart rate, activity, and sleep metrics measured from wearable devices was able to identify 63% of COVID-19 cases before symptoms, further demonstrating the promise of this approach.^6,7^

Our group launched the Warrior Watch Study, which employed a custom smartphone app to remotely monitor health care workers (HCWs) throughout the Mount Sinai Health System.^8^ This app delivered surveys to the subject’s iPhones and enabled passive collection of Apple Watch data. We previously demonstrated that significant changes in heart rate variability (HRV), the small differences in time between each heartbeat that reflect autonomic nervous system (ANS) function, collected from the Apple Watch, occurred up to 7 days before a COVID-19 diagnosis.^8,9^ Building on these observations, our primary aim was to determine the feasibility to train and validate machine learning approaches combining HRV measurements with resting heart rate (RHR) metrics to predict COVID-19 before diagnosis via nasal polymerase chain reaction (PCR).

## Methods

### Study Design

We recruited HCWs for this prospective observational study from seven hospitals in New York City (The Mount Sinai Hospital, Morningside Hospital, Mount Sinai West, Mount Sinai Beth Israel, Mount Sinai Queens, New York Eye, and Ear Infirmary, Mount Sinai Brooklyn).^8^ Subjects were ≥18 years, employees at one of these hospitals, had at least an iPhone series 6, and were willing to wear an Apple Watch Series 4 or higher. Underlying autoimmune or inflammatory diseases, as well as medications known to interfere with ANS function, were exclusionary. The study was approved by the Mount Sinai Hospital Institutional Review Board, and all subjects provided informed consent prior to enrollment.

### Study Procedures

Subjects downloaded the Warrior Watch Study app, signed the electronic consent, and completed baseline demographic questionnaires. Prior COVID-19 diagnosis, medical history, and occupation classification within the hospital were collected via in-app assessments. Subjects completed daily surveys to report any COVID-19 related symptoms, symptom severity, the results for any SARS-CoV-2 nasal PCR tests, and SARS-CoV-2 antibody test results. A positive diagnosis was defined as a self-reported positive SARS-CoV-2 nasal PCR test. Subjects were asked to wear the Apple Watch for at least 8 hours per day (**Figure 1a**).

**Figure 1.**
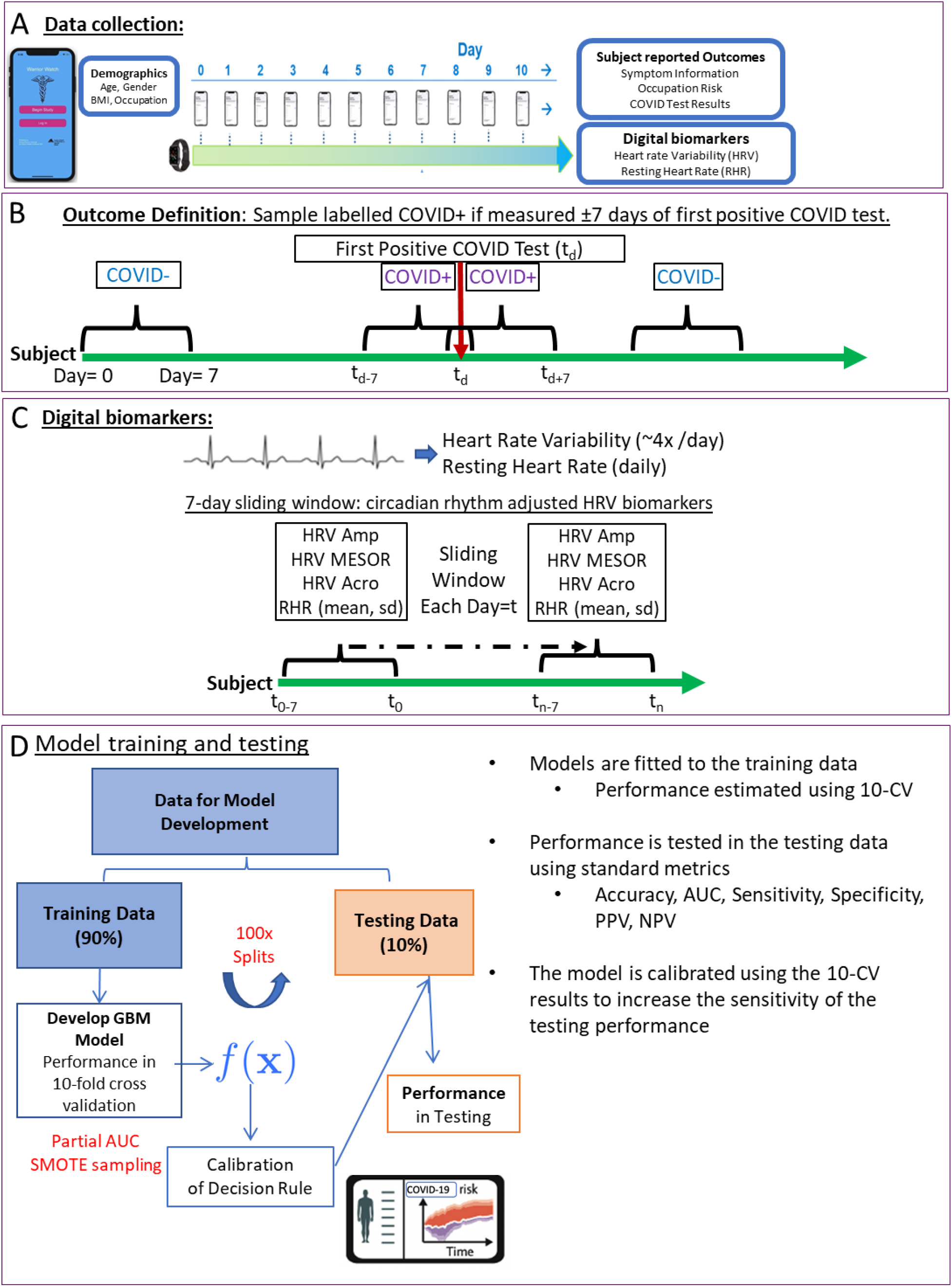
General Strategy for training and testing statistical classifiers. Diagram illustrating the general strategy for developing the statistical classifier. (A) Subjects wore smartwatches that collect measurements of HRV and RHR. Subjects answer daily surveys to provide health outcomes including COVID test results. (B) Each day each subject is labelled as either; COVID+ if observation was made within ±7 days of the patients first positive COVID-19 test, otherwise the observation is labelled COVID-. (C) HRV measurements were too sparse to estimate HRV COSINOR parameters (MESOR, Amplitude, Acrophase) for each day, thus, we estimated smoothed parameters using a 7-day sliding window. RHR (mean, sd, min, max) was also estimated over this window. (D) The data was split into 100 training and testing sets, models were fit to the training data and performance was estimated using 10-fold CV. 10-CV predictions were used define a decision rule that increases sensitivity, this decision rule was applied to the predictions in the testing data to get the final performance.

### Wearable Device

Subjects wore an Apple Watch Series 4 or higher, which are commercially available wearable devices that connect via Bluetooth to participants’ iPhones. The Apple Watch uses infrared and visible-light light-emitting diodes and photodiodes that act as a photoplethysmogram generating time series peaks from each heartbeat.^10^ There is a moving average window during which heart rate measurements are calculated while the device is worn. HRV is automatically calculated in ultra-short 60 second recording periods as the standard deviation of the inter-beat interval of normal sinus beats (SDNN), a time-domain index.^9^ SDNN reflects sympathetic and parasympathetic nervous system activity. The Warrior Watch Study app collects the generated SDNN and heart rate measurements at survey completion.

### Data handling, model development and statistical analysis

Our primary analysis consisted of measurements of HRV. HRV follows a circadian pattern that can be characterized by three parameters, namely the MESOR (M: the mean HRV during the day), amplitude (A: maximum HRV during the day), and the acrophase (Ψ: describing when the maximum occurs).^8^ We previously developed a mixed-effects COSINOR model to compare HRV circadian patterns at the group level and show that changes in those parameters were associated with infection.^8^ Given these findings, daily measurements of HRV were incorporated as potential diagnostic biomarkers for our machine-learning approach.

HRV measurements for each day were sparse and were not taken at regular intervals. Thus, daily estimates of HRV COSINOR parameters M, A and Ψ could not be calculated. Due to this limitation, we estimated the daily HRV parameters for each subject and day (t_n_) using HRV data from a seven-day sliding window (t_n_ – t_n-6_), thereby creating daily smoothed estimates reflecting changes in the last 7 days (**Figure 1b**). To aid the optimization procedures, each subject’s initial estimates are obtained using the first two weeks of data from each subject fitted to a mixed-effect COSINOR model with A, M, and Ψ as random effects.^8^ From this model, the subject-specific baseline A, M, and Ψ is derived and used to initialize the iterative 7-day smoothed estimates within each patient. If the number of days in the 7-day window was < 3, the window was expanded to 14 days (t_n-14_). In rare cases, no data was available over 14-days, and parameters were imputed using the Last Observation Carried Forward (LOCF) imputation method. During each window, we also measured the maximum, minimum, mean, and standard deviation (SD) of the RHR. For each day and subject, there were a total of 8 digital biomarkers used to develop our predictive models: HRV-amplitude, HRV-MESOR, HRV-acrophase, daily RHR, RHR-max, RHR-min, RHR-sd, RHR-mean, and 3 demographic variables known to impact HRV-BMI, age, and gender.^11^

Data was split into independent training and testing sets, ensuring that observations were taken on chronologically similar days (e.g., Day 6 and Day 7), for the same subject, were in the same set. A sampling procedure was employed that ensured that observations with proximity in time (±4 days), for the same subject, did not appear in both training and testing sets. This procedure was created 100 training and testing sets, containing 90% and 10% of the data respectively. Care was also taken to ensure that the prevalence of COVID-19 positive (COVID+) diagnoses in each set was similar to the prevalence of the full data set.

Machine learning model training and evaluation were performed using *caret* and *pROC* packages, with tuning parameters estimated using 10-fold cross-validation (10-CV). To safeguard against biases induced by the low prevalence of COVID+ samples, we considered several sampling methods to balance the data during 10-CV, ultimately using SMOTE (synthetic minority over-sampling technique) which had the most favorable 10-CV performance.^12^ Models were trained on each of the 100 training sets, and their performance (auROC, partial-auROC, accuracy, positive predictive value (PPV), negative predictive value (NPV), sensitivity, specificity, balanced accuracy) was assessed on the corresponding testing set and presented as mean with 95% CI. The sensitivity of the diagnostic algorithm was prioritized since the application of wearable devices as a non-invasive screening modality would be to prompt a confirmatory PCR test. Our models were trained to maximize partial-auROC (sensitivity boundary of >75%), with tuning parameters estimated using 10-fold cross-validation (CV). When exploring the training data, 10-CV performance for several different machine-learning algorithms was assessed (gradient-boosting machines, elastic-net, partial least squares, support vector machines and random forests). However, a gradient boosting machine model (GBM) was selected as the best performing and was used to develop our statistical classifier (**Supplementary Table 1**).

When calibrating the model, the 10-CV predictions were used to optimize the probability threshold such that the sensitivity was above >98%. The average value of this probability threshold, over all 100 iterations, was then used to define the final decision rule where cases with a predicted probability above this threshold were considered COVID+. We used a previously described method to estimate each feature’s relative influence/importance in the model, over all 100 training sets.^13^ All analyses were performed by R, version 4.0.2, including the *caret* and *pROC* packages.^14,15^

## Results

### Study population

Four hundred and seven HCWs were enrolled between April 29^th^, 2020, and March 2^nd^, 2021 (**Table 1**). The mean age of participants at enrollment was 38 years (SD 9.8), and 34.2% were men. A positive SARS-CoV-2 nasal PCR was reported by 12.0% (49/407) of participants during follow-up (**Figure 1c**). The median follow-up time was 73 days (range, 3-253 days) for a total of 28,528 days of observations. A median of 4 HRV samples were collected at varying times per participant per day, and daily measures of RHR.

**Table 1.** Baseline characteristics of study participants.

|  | Total Cohort<br>(n=407) | Never COVID-19<br>Positive<br>(n=358) | COVID-19<br>Positive<br>(n=49) | P- value |
| --- | --- | --- | --- | --- |
| Age, mean (SD) | 37.9 (9.82) | 37.9 (9.73) | 37.3 (10.55) | 0.65 |
| Body Mass Index, mean (SD) | 25.7 (5.47) | 25.7 (5.50) | 25.8 (5.31) | 0.91 |
| Male Gender (%) | 139 (34.2) | 128 (35.8) | 11 (22.4) | 0.09 |
| Race (%) |  |  |  | 0.07 |
| Asian | 111 (27.3) | 104 (29.1) | 7 (14.3) |  |
| Black | 43 (10.6) | 40 (11.2) | 3 (6.1) |  |
| Hispanic/Latino | 71 (17.4) | 58 (16.2) | 13 (26.5) |  |
| Other | 23 (5.7) | 21 (5.9) | 2 (4.1) |  |
| White | 159 (39.1) | 135 (37.7) | 24 (49.0) |  |
| Baseline Negative SARS-CoV-2<br>Nasal PCR (%) | 382 (93.9) | 346 (96.6) | 36 (73.5) | <0.001 |
| Baseline Negative SARS-CoV-2<br>serum antibody (%) | 367 (90.2) | 325 (90.8) | 42 (85.7) | 0.39 |
| Baseline Smoking Status (%) |  |  |  | 0.61 |
| Never/Rarely smoker | 343 (84.3) | 300 (83.8) | 43 (87.8) |  |
| Current/Past smoker | 64 (15.7) | 58 (16.2) | 6 (12.2) |  |

COVID-19, coronavirus disease 2019; PCR, polymerase chain reaction; SD, standard deviation

### Performance in training and 10-CV, and model calibration

Given the low prevalence of COVID+ observations (<1% of all daily observations were COVID+), and to avoid biased performance metrics resulting from a single split, the data was split into 100 training (including ∼90% of the data) and testing (∼10%) sets, using a strategy that guarantees independence between testing and training sets. This procedure produced robust estimates of the model performance in the testing set as well as 95% CI (**Figure 1D**). The 10-CV performance of several different machine-learning methods was explored (**Supplementary Table 1**). This analysis revealed that the non-linear methods (AUC>98%), GBM and random forest (RF), outperformed all linear methods (AUC<66%), suggesting a non-linear relationship between HRV and SARS-CoV-2 infection. Although the RF model had a higher AUC than GBM, the training performance of RF was consistently high and demonstrated a larger discrepancy between training and 10-CV sensitivity (**Supplementary Figure 1**). Given this overfitting of the data we used GBM to develop our final classifier.

ROC curves calculated for GBM using all training and 10-CV samples show a high AUC (>98%) (**Figure 2a-b**). The sensitivity and specificity were >95% in the training data (**Supplementary Table 2**). However, the sensitivity in 10-CV was comparatively lower (∼91%) (**Figure 2a**), likely due to the low prevalence of COVID+ examples in the training data and biasing the model to predict COVID-outcomes, suggesting that sensitivity estimates in the training data are an overestimate.

**Figure 2:**
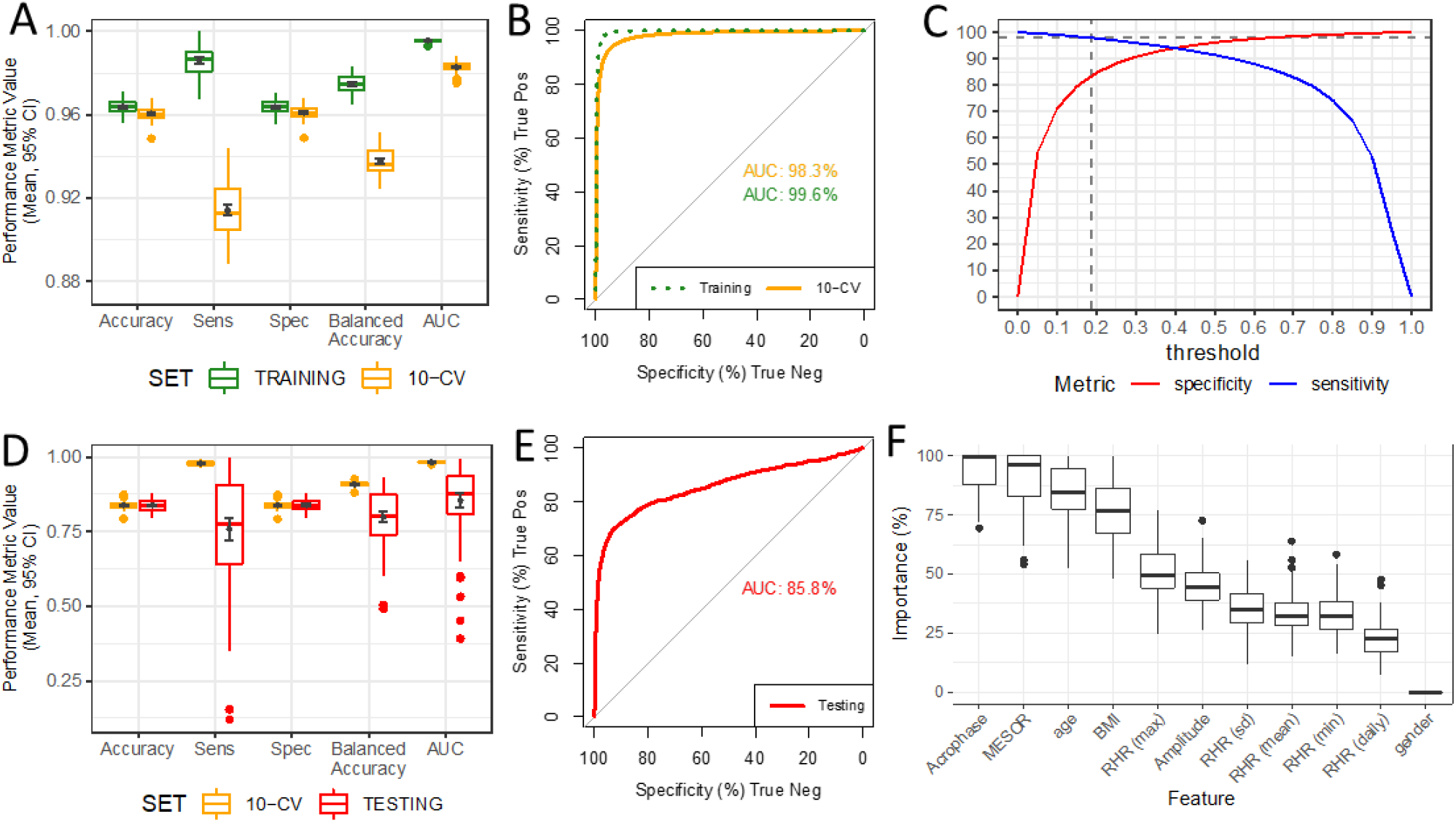
Model performance in training and testing data. (A) Boxplots show distribution of performance metrics in training data and 10-fold CV over all 50 training sets. (B) ROC curve and AUC over all training and 10-fold CV samples. (C) Plot shows specificity (red) and sensitivity (blue) at different response thresholds for all 10-fold CV samples, a threshold ∼0.3 achieved a sensitivity of 95% and a specificity of 55%. (D) ROC curve and AUC over all testing samples. (E) Distribution of accuracy, specificity and sensitivity over all 50 10-fold CV and test sets using the 0.3 threshold decision rule. (F) Distribution of feature importance metrics estimated from the models trained on each of the 50 training sets, a higher importance indicates that the features made a higher contribution to prediction.

We calibrated the final decision rule to guarantee high sensitivity, as a wearable device-based algorithm would be utilized for screening (**Table 2**). This calibrated decision rule increases the true positive rate by allowing for a larger rate of false-positive results. To keep the testing performance unbiased, we used the training data to optimize the decision rule to guarantee a sensitivity >98% (**Figure 2c**). This optimal decision rule was 0.19 (**Figure 2c**) and produced an average 10-CV Accuracy (**Figure 2d**) of 84% (CI ±∼0.3%), with 98% sensitivity and 84% specificity, thus indicating a specificity loss of 12%, for a 7% gain in sensitivity compared to the standard 0.5 decision threshold. When the calibrated diagnostic rule was applied to testing data, an AUC >85% (**Figure 2d-e**) was archived (median= 88%). Accuracy and specificity were 84% (CI ±∼0.4%) (**Figure 2d**). The mean sensitivity was 76% (CI ±∼4%).

**Table 2.**
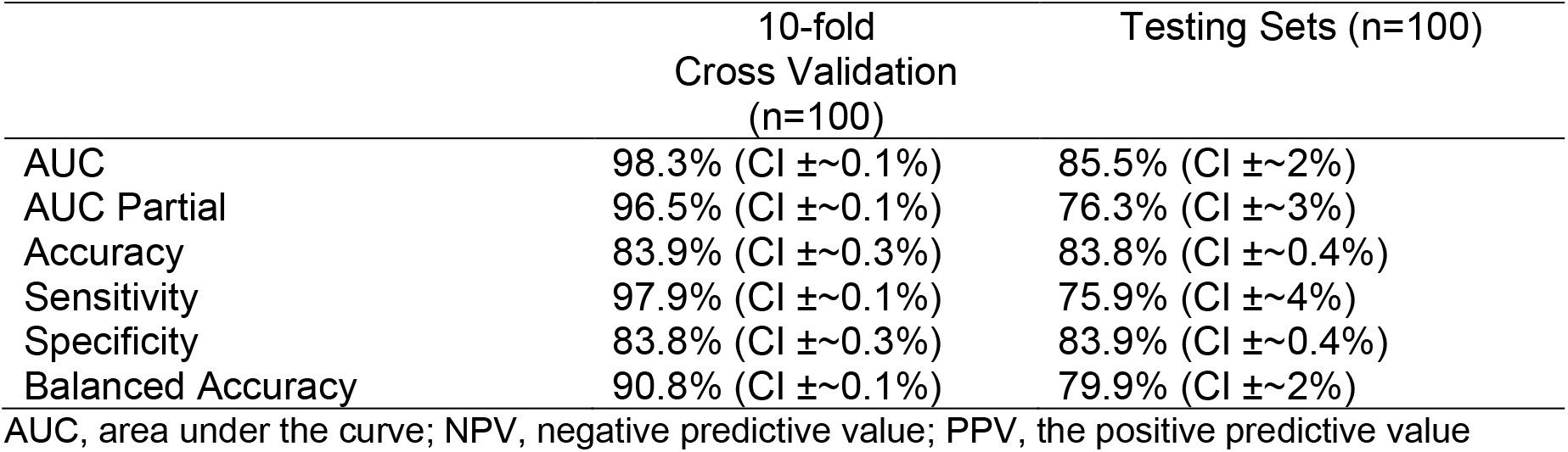
Performance summary of the gradient boosting machine learning model in 10- fold cross-validation and testing sets after calibration.

### Feature importance and interpretation

The four most important/influential predictors were HRV acrophase, HRV MESOR, age and BMI (**Figure 2f**), with median importance >75%. RHR metrics (maximum, minimum, SD, mean) as well as HRV amplitude, were less influential (median importance 25-50%). Gender had importance equal to 0 in most models. To visualize the relationship between feature values and model prediction, we selected the 9 patients for which the model was best able to predict COVID-19 (AUC>98.9% 10-CV), and plotted the acrophase, amplitude, MESOR and max RHR, as well as the predicted probability, for each day (**Figure 3**). This analysis revealed a complex relationship between HRV parameters and SARS-CoV-2 infection. It was notable that, for most subjects, the predicted probability increased when HRV amplitude decreased, which is consistent with our previously published analysis.^8^

**Figure 3:**
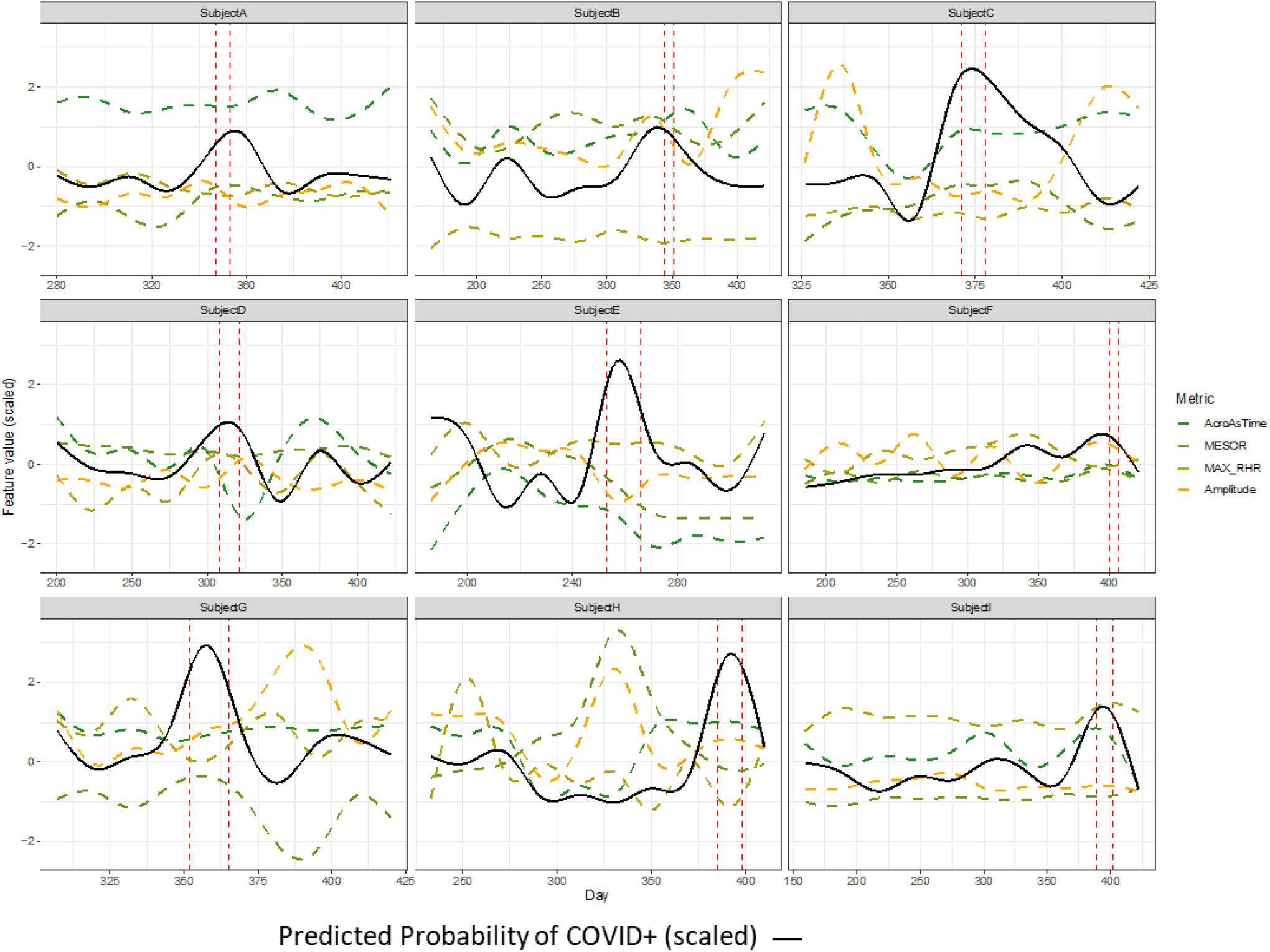
Changes in HRV parameters and model predictions over time. Line plots show daily measurements of HRV parameters (Acrophase, MESOR, Amplitude), and Maximum RHR, as well as the probability of infection (black line) predicted by the model. All values are centered, scaled and smoothed to facilitate comparison. Daily measurements for 9 subjects are shown, predictions for each of these 9 subjects all had AUC>98.9% in 10-CV. Vertical red-dashed lines indicate the infection window for each patient.

## Discussion

Our results demonstrate that a machine learning approach applied to the physiological metrics measured by a wearable device reliably identifies and predicts SARS-CoV-2 infections. This highlights the potential utility of assessing individual changes in passively collected physiological data from wearable devices to facilitate the management of the COVID-19 pandemic.

Infections alter physiological metrics differentiating infected and uninfected states. Changes in vital signs in the setting of infection, including increased heart rate, elevated respiratory rate, and altered body temperature, have been well described.^16,17^ In addition to these traditional physiological metrics, ANS function, measured by HRV, is altered during illness. Several small studies have shown that changes in HRV can identify and predict infections.^18,19^ Building on these observations and the growing capabilities of wearable technology, wearable devices have been increasingly explored in the setting of infection. They provide a unique means to measure physiological parameters and offer an advantage over periodic assessments in the clinical setting by collecting real-time continuous measurements.^20^ This approach can identify trends in individual physiological outputs. On a population level, retrospective analysis of physical activity and heart rate data collected from Fitbits was shown to improve influenza-like illness predictions.^21^ This approach applied to an individual level was explored during the COVID-19 pandemic.

SARS-CoV-2 alters physiological metrics commonly measured by wearable devices.^22^ Quer et al. and colleagues collected symptom data and physiological metrics from smartwatches. They found that while resting heart rate could not discriminate SARS-CoV-2 infections from negative cases (AUC of 0.52) when combined with sleep, activity, and symptom-based data, the AUC increased to 0.80. They demonstrated that the addition of wearable-based data significantly improved the ability of symptoms alone to discriminate between those positive or negative for COVID-19.^6^ Similarly, Mishra et al. demonstrated that heart rate, physical activity, and sleep time were collected from wearable devices could detect COVID-19. They found that 26 of the 32 COVID-19 positive subjects in their cohort had significant alterations of these metrics before diagnosis or symptom development and that 63% of cases could be detected before symptom onset.^7^

Our group previously demonstrated that changes in the circadian pattern of HRV were associated with a COVID-19 diagnosis.^8^ We demonstrated that significant changes, particularly in the amplitude of SDNN, were observed over the 7 days before diagnosis. Based on this observation, we built a machine learning algorithm that incorporated HRV circadian rhythm, RHR parameters, and demographic characteristics that can easily be collected from wearable device users. We trained a predictive model and then demonstrated the ability to accurately predict COVID-19 status in new data with relatively high sensitivity (76%) and specificity (84%), compared to the current gold standard of SARS-CoV-2 nasal PCR testing. This model’s high sensitivity and the minimal demographic data required lends itself to easy deployment. Our model has an advantage over prior publications evaluating the relationship between wearable-based data and a COVID-19 diagnosis, in that we trained a predictive model and then demonstrated its accuracy in predicting COVID-19 status in new data.^6-8^

There are several limitations to our study. First, HRV was collected sporadically by the Apple Watch. We employed statistical modeling to account for this. However, a denser data set using continuous data would likely further improve our predictions. Second, the model we employed used a 7-day smoothing approach. This approach observed infection-induced changes in HRV later than if HRV was estimated using a single-day method. Thus, the approach we employed is conservative. An additional limitation is that the Apple Watch provides HRV measurements only in the SDDN time domain. This limits assessments between other types of HRV measurements and COVID-19 outcomes. Additionally, other factors might impact HRV, which we were not able to capture and control for in the analysis. Furthermore, we were not routinely checking for SARs-CoV-2 infections and relied on subjects reporting a COVID-19 diagnosis. Therefore, infections could have occurred that are not accounted. Lastly, we did not externally validate our machine learning algorithm in another cohort.

## Conclusion

We demonstrate that a machine learning algorithm combining circadian features of HRV with features of resting heart rate derived from the Apple Watch achieves high sensitivity and specificity in predicting the development of COVID-19. While further validation is necessary, this non-invasive and passive modality may be helpful to monitor large numbers of people for possible infection with SARS-CoV-2 and help direct testing toward high-risk individuals.

## Data Availability

Data used in the present study includes health care worker data that is not available upon request.

---

**Supplementary Table 1.** 10-CV performance of multiple machine learning methods.

| Machine Learning Method | Area Under Receiver Operating Characteristic | Area Under Partial Receiver Operating Characteristic (Sensitivity>0.75) | Sensitivity | Specificity |
| --- | --- | --- | --- | --- |
| Random Forest | 0.99 | 0.98 | 0.93 | 0.99 |
| Gradient Boosting Machines | 0.98 | 0.97 | 0.92 | 0.96 |
| Elastic Net Regularization | 0.67 | 0.61 | 0.44 | 0.78 |
| Support-Vector Machine (Linear) | 0.66 | 0.59 | 0.40 | 0.80 |
| Partial Least Squares | 0.65 | 0.61 | 0.36 | 0.79 |

Roc, receiver operating characteristic

**Supplementary Table 2.** Performance summary of the gradient boosting machine learning model in training sets and 10- fold cross-validation.

|  | Training Sets (n=100) | 10-fold Cross Validation (n=100) |
| --- | --- | --- |
| AUC | 99.6% (CI ±~0.1%) | 98.3% (CI ±~0.1%) |
| AUC Partial | 99.3% (CI ±~0.1%) | 96.5% (CI ±~0.1%) |
| Accuracy | 96.4% (CI ±~0.1%) | 96.1% (CI ±~0.1%) |
| Sensitivity | 98.6% (CI ±~0.2%) | 91.4% (CI ±~0.2%) |
| Specificity | 96.4% (CI ±~0.1%) | 96.1% (CI ±~0.1%) |
| Balanced Accuracy | 97.4% (CI ±~0.1%) | 93.7% (CI ±~0.1%) |

AUC, area under the curve; NPV, negative predictive value; PPV, the positive predictive value

**Supplementary Figure 1:**
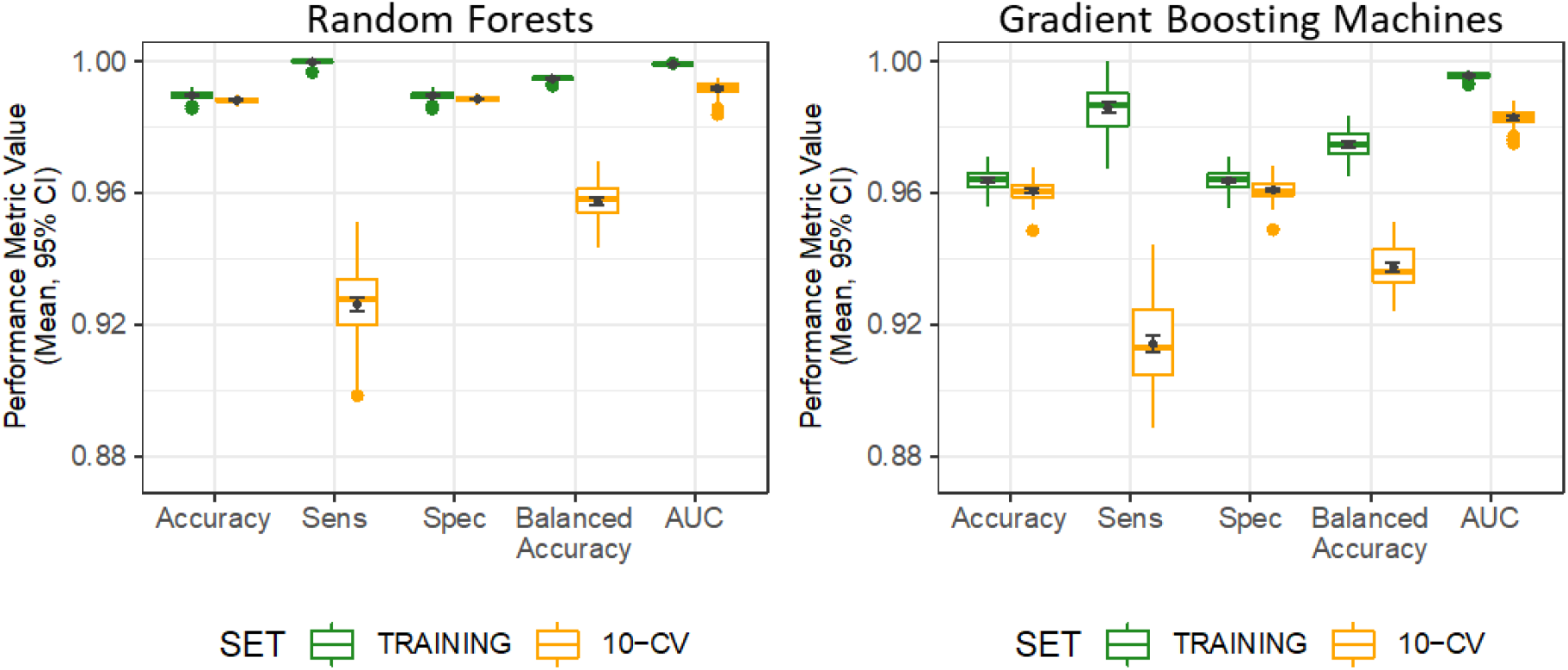
Training a 10-CV performance for RF and GBM models. Box plots show distribution of performance metrics in training (green) and 10-CV (yellow) for random forests and gradient boosting models, over 100 training sets. Mean +/- 95% confidence intervals are shown in black.

